# Collider bias undermines our understanding of COVID-19 disease risk and severity

**DOI:** 10.1101/2020.05.04.20090506

**Authors:** Gareth J Griffith, Tim T Morris, Matt Tudball, Annie Herbert, Giulia Mancano, Lindsey Pike, Gemma C Sharp, Tom M Palmer, George Davey Smith, Kate Tilling, Luisa Zuccolo, Neil M Davies, Gibran Hemani

**Author notes:** Contributed equally (Gareth J Griffith, Tim T Morris, Matt Tudball, Annie Herbert, Giulia Mancano, Gibran Hemani).

## Abstract

Observational data on COVID-19 including hypothesised risk factors for infection and progression are accruing rapidly, often from non-random sampling such as hospital admissions, targeted testing or voluntary participation. Here, we highlight the challenge of interpreting observational evidence from such samples of the population, which may be affected by collider bias. We illustrate these issues using data from the UK Biobank in which individuals tested for COVID-19 are highly selected for a wide range of genetic, behavioural, cardiovascular, demographic, and anthropometric traits. We discuss the sampling mechanisms that leave aetiological studies of COVID-19 infection and progression particularly susceptible to collider bias. We also describe several tools and strategies that could help mitigate the effects of collider bias in extant studies of COVID-19 and make available a web app for performing sensitivity analyses. While bias due to non-random sampling should be explored in existing studies, the optimal way to mitigate the problem is to use appropriate sampling strategies at the study design stage.

## Introduction

Government health organisations, researchers and private companies, amongst others, are generating data on the COVID-19 status of millions of people, along with measures of health and behaviour, for the purpose of using these samples to understand the *risk factors* (see **Box 1** for scope) relevant to the disease in the general population. Numerous studies have reported risk factors associated with COVID-19 infection and subsequent disease severity, such as age, sex, occupation, smoking, and ACE-inhibitor use (1-10). But if we are to make reliable inference about the causes of infection and severity, we need to be aware that there are serious limitations to such observational data. Of particular importance to understanding the aetiology of COVID-19 or developing predictors for infection or severity is the problem of collider bias (sometimes referred to as selection bias, sampling bias, ascertainment bias). Emerging datasets relating to COVID-19 may be particularly susceptible to this issue, having serious implications for the reliability of causal inference and generalisability of predictors.

A collider is a variable that is influenced by two other variables of interest (they “collide” in a Direct Acyclic Graph) and can be considered the contrast of a confounder, which influences other variables (**Figure 1A**). Collider bias can be unintuitive and its implications highly context-specific, but several illustrative examples have been published to aid with understanding this issue (11-13). Consider the situation where we want to test whether a hypothesised risk factor (e.g. age) causes an outcome (e.g. contracting COVID-19). If the hypothesised risk factor and the outcome each influence a third variable, conditioning on that third variable will induce an artifactual association between the risk factor and the outcome even if the risk factor does not cause the outcome (**Figure 1, 2**). If this third variable is participation in a study (i.e. an observation being present in the dataset), then analysis of the study sample will automatically condition on the collider of participation. Collider bias can induce associations where there is no true causal effect in the general population, attenuate or inflate true causal effects, or reverse the sign of true causal effects.

**Figure 1:**
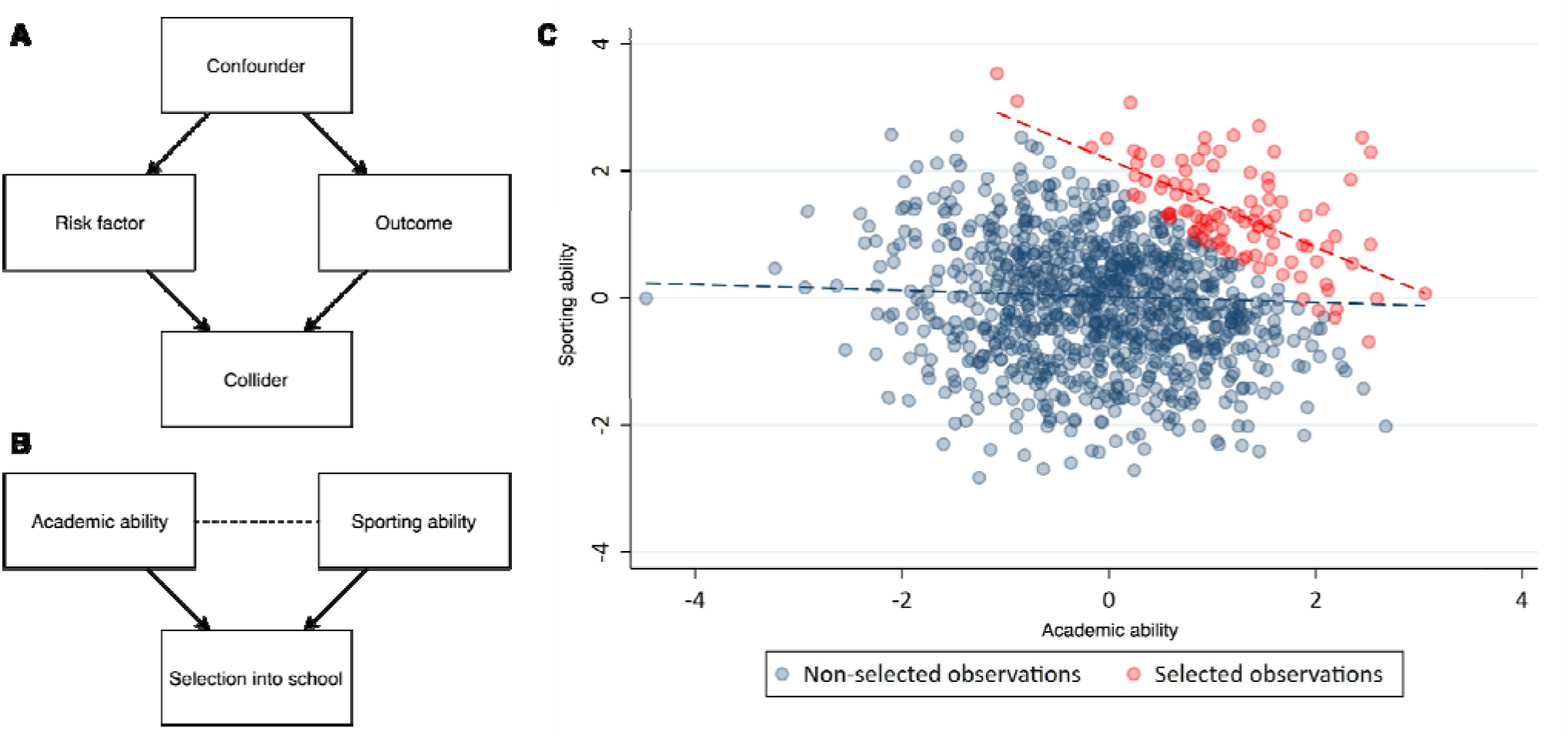
Illustrative example of collider bias. A) A directed acyclic graph (DAG) illustrating definitions of a collider and a confounder with respect to a model analysing the association between an hypothesised risk factor and an outcome. Directed arrows indicate causal effects. B) A DAG representing the simple example presented in the main text. Academic ability and sporting ability each influence selection into a prestigious school. The dotted line indicates an induced correlation, as shown in (C), these traits are not correlated in the general population, but because they are selected for enrollment they become correlated when analysing only the selected individuals.

**Figure 2:**
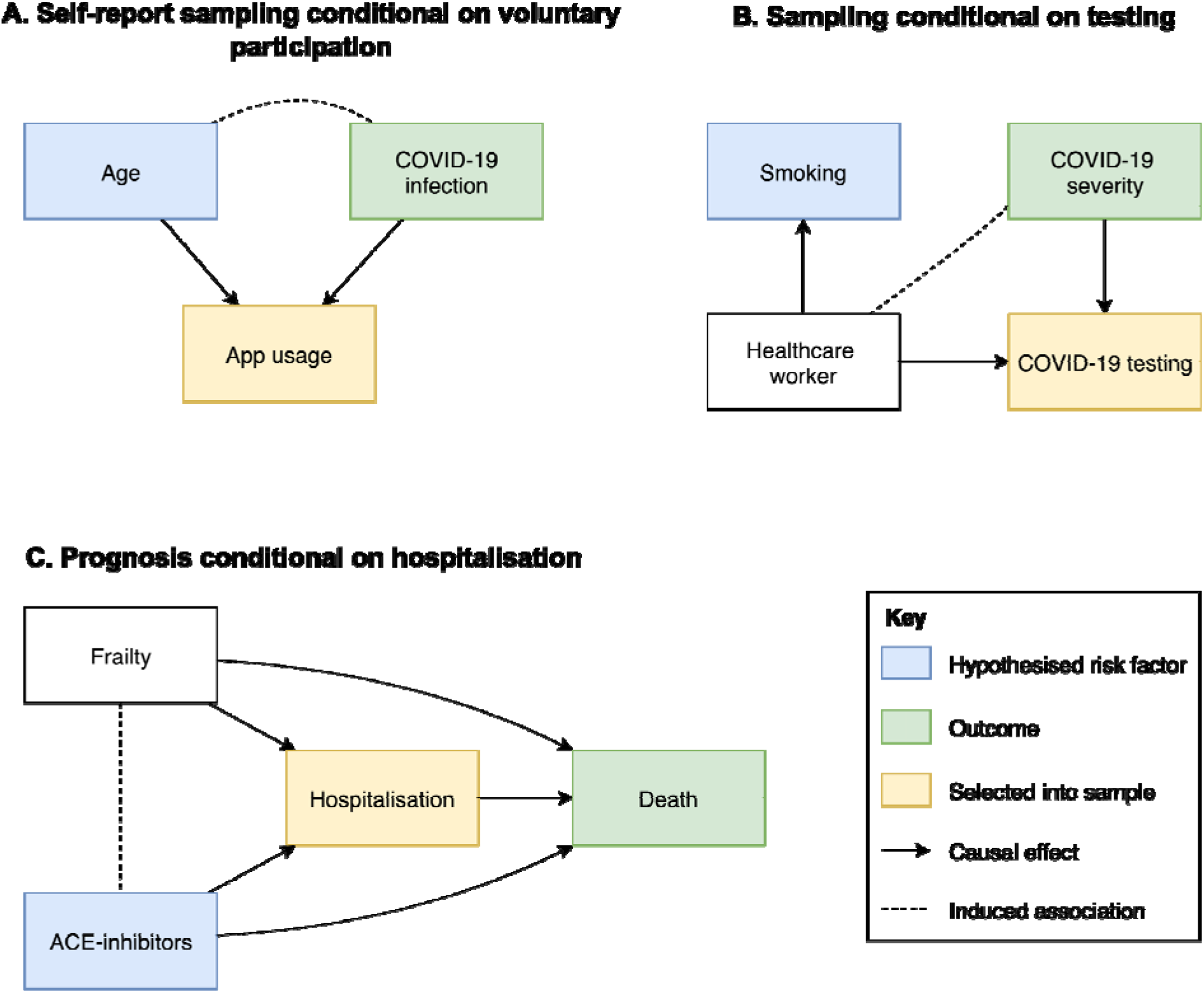
Collider bias induced by conditioning on a collider in three scenarios relating to COVID-19 analysis. These are simplified Directed Acyclic Diagrams where only the main variables of interest have been represented for sake of illustrating collider bias scenarios. All assume no unspecified confounding or other biases. Rectangles represent observed variables and solid directed arrows represent causal effects. The dashed line represents an induced association when conditioning on the collider, which in these scenarios are variables that indicate whether an individual is selected into the sample. (A) When some hypothesised risk factor (e.g. age) and outcome (e.g. COVID-19 infection) each associate with sample selection (e.g. voluntary data collection via mobile-phone apps), the hypothesised risk factor and outcome will be associated within the sample. The presence and direction of these biases are model dependent; where causes are supra-multiplicative they will be positively associated in the sample; where they are sub-multiplicative they will be negatively correlated; and where they are exactly multiplicative they will remain unassociated. We extend this scenario in (B) where the association between the hypothesised risk factor and the collider does not need to be causal. (C) When inferring the influence of some hypothesised risk factor on mortality, in an unselected sample the risk factor for infection is a causal factor for death (mediated by COVID-19 infection). However, if analysed only amongst individuals who are known to have COVID-19 (i.e. we condition on the COVID-19 infection variable) then the risk factor for infection will appear to be associated with any other variable that influences both infection and progression. In many circumstances this can lead to a risk factor for disease onset that appears to be protective for disease progression. Each of these scenarios represent those described in the main text.

In the context of this paper, conditioning upon the third variable can mean examining the effect of the risk factor in only the subset of individuals with that particular characteristic (e.g. only analysing disease cases for disease progression), or non-randomly selecting samples from the target population. Two intuitive examples of collider bias are laid out below.

**Example 1:** Suppose that sporting ability and academic ability are both normally distributed and independent in the population. That is, sporting ability has no influence on academic ability, and vice versa. Let’s now suppose that a prestigious, highly selective school chooses to enrol children who have high sporting or academic ability (**Figure 1B**). For ease, assume that the school has sufficient capacity to enrol the top 10% of pupils from the general population, based on their combined sporting and academic scores. Because sporting and academic ability are independent in the general population and we have selected on the top of these distributions, enrolled pupils are likely to be either sporting or academic, and unlikely to be both. This induces a negative correlation in our school despite there being no relationship in the general population (**Figure 1C**). If we analyse sporting and academic ability in the school, we will conclude that the two are inversely related. It is clear that analysis within this selected sample will generate unreliable causal inference, and unreliable predictors to be applied to the general population.

**Example 2:** Suppose we want to test the hypothesis that being a health worker is a risk factor for severe COVID-19 symptoms. Our target population for hypothesis is all adults in the general population. However, our study sample will be restricted only to those who are tested for active COVID-19 infection. If we take the UK as an example (until late April 2020), the majority of tests were performed either on health workers, or members of the general public who had symptoms severe enough to require hospitalisation. In this testing environment, our sample of participants will be selected for both the hypothesised risk factor (being a healthcare worker) and the outcome of interest (severe symptoms). In this strata of the population, healthcare workers will generally appear to have relatively low severity (inducing a negative observational association, **Figure 2B**). In reality, there are occupational hazards relating to infectious disease of being a healthcare worker, and the true causal effect is likely to be in the opposite direction, as healthcare workers are likely exposed to higher viral loads.

In this paper, we discuss why collider bias should be of particular concern to observational studies of COVID-19, and show how sample selection can lead to dramatic biases. We then go on to describe the approaches that are available to explore and mitigate this problem.

## Why observational COVID-19 research is particularly susceptible to collider bias

Though unquestionably valuable, observational datasets can be something of a black box because the associations to which they give rise can be due to many different mechanisms. Suppose we wish to draw inferences that can be generalised to a wider population such as the UK (the population). To conduct our observational study, we must first define a group of people that we wish to sample (the target population) who are representative of the target population. The members of the target population who respond to the invitation and participate in the study form the study sample. If individual characteristics cause people to be more likely to respond to an invitation to participate in the study, the study sample will not be representative of the target population. To give context on how serious a problem collider bias can be, there is a continuing debate in the literature about the extent to which it is appropriate to adjust for covariates in observational associations (14-17). If we assume that a given covariate influences both the hypothesised risk factor and the outcome, it is appropriate to condition on that covariate to remove bias induced by the confounding structure. However, if the covariate is a common consequence rather than a common cause, then we risk inducing, rather than reducing bias (18). That is, collider bias can also be introduced when making statistical adjustments for variables that lie on the causal pathway between exposure and outcome.

*A priori* knowledge of what the hidden causal structure truly is can be hard to deduce, and it is appropriate to treat collider bias with a similar level of caution to confounding bias.

There are multiple ways in which data are being collected on COVID-19, and they can introduce unintentional conditioning in the selected sample in various ways. The characteristics of participants recruited are related to a range of factors including policy decisions, cost limitations, technological access, and testing methods. It is also widely acknowledged that the true prevalence of disease in the population remains unknown (19). Here we describe the forms of data collection for COVID-19 and then go on to detail the circumstances surrounding COVID-19 that make its analysis susceptible to collider bias.

### COVID-19 sampling strategies and case definitions

#### Sampling conditional on voluntary participation (Case definition: probable COVID-19, Figure 2A)

Probable COVID-19 status can be determined through studies that require voluntary participation. These may include, for example, surveys conducted by existing cohort and longitudinal studies (20,21), data linkage to administrative records is also available in some cohort studies such as the UK Biobank (22), or mobile phone based app programmes (23,24). Participation in scientific studies has been shown to be strongly non-random (e.g. participants are disproportionately likely to be highly educated, health conscious, and non-smokers), so the volunteers in these samples are likely to differ substantially from the general population (25-27). See **Box 2** for a vignette on how one study (24) explored collider bias in this context.

#### Sampling conditional on being tested for active COVID-19 infection (Case definition: positive test for COVID-19, Figure 2B)

Polymerase chain reaction (PCR) antigen tests are used to confirm a suspected (currently active) COVID-19 infection. Studies that aim to determine the risk factors for confirmed current COVID-19 infection therefore rely on participants having received a COVID-19 antigen test (hereafter for simplicity: COVID-19 test or test). Unless a random sample or the entire population are tested, these studies do not provide an unbiased estimate of active COVID-19 infection prevalence in the general population. As testing is a resource limited endeavour, different countries have been using different (pragmatic) strategies for prioritising testing, including on the basis of characteristics such as occupation, symptom presentation and perceived risk. See **Box 3** for an investigation into the extent to which testing is non-random with respect to a range of measurable potential risk factors, using the recently released COVID-19 test data in the UK-Biobank.

Note that the example shown in **Figure 2B**, It is possible to induce a correlation between two variables that are not directly influencing sample selection. If the two variables that influence sample selection themselves influence other traits, those other traits will be correlated.

#### Sampling conditional on having a positive test for active COVID-19 infection (Case definition: severe COVID-19 symptoms, Figure 2B)

Studies that aim to determine the risk factors for severity of confirmed current COVID-19 infection therefore rely on participants having received a COVID-19 antigen test (hereafter for simplicity: COVID-19 test or test), and that the result of the test was positive. As above, testing is unlikely to be random, and conditioning on the positive result will also mean bias can be induced by all factors causing infection, as well as those causing increased likelihood of testing.

#### Sampling conditional on hospitalisation (Case definition: COVID-19 infection)

An important source of data collection is from existing patients or hospital records. Several studies have emerged which make causal inference from such selected samples (8,9,28). COVID-19 infection influences hospitalisation, as do a large number of other health conditions. By analysing only hospitalised samples, anything that influences hospitalisation will become negatively associated with COVID-19 infection (appearing protective).

#### Prognosis and mortality sampling conditional on hospitalisation (Case definition: COVID-19 death, Figure 2C)

Many studies have started analysing the influences on disease progression once individuals are infected, or infected and then admitted to hospital (i.e. the factors that influence survival). Such datasets necessarily condition upon a positive test. **Figure 2C** illustrates how this so-called ‘index event bias’ is a special case of collider bias (29-31). If we accept that COVID-19 increases mortality, and there are risk factors for infection of COVID-19, then in a representative sample of the target population, any cause of infection would also exert a causal influence on mortality, mediated by infection. However, once we condition on being infected, all factors for infection become correlated with each other. If some of those factors influence both infection and progression then the association between a factor for infection and death in the selected sample will be biased. This could lead to factors that increase risk of infection falsely appearing to be protective for severe progression (1,32). An example of this relevant to COVID-19 is discussed in **Box 2**. How different directions of selective sampling influence the direction of bias is discussed in **Figures 2 and 3**.

**Figure 3:**
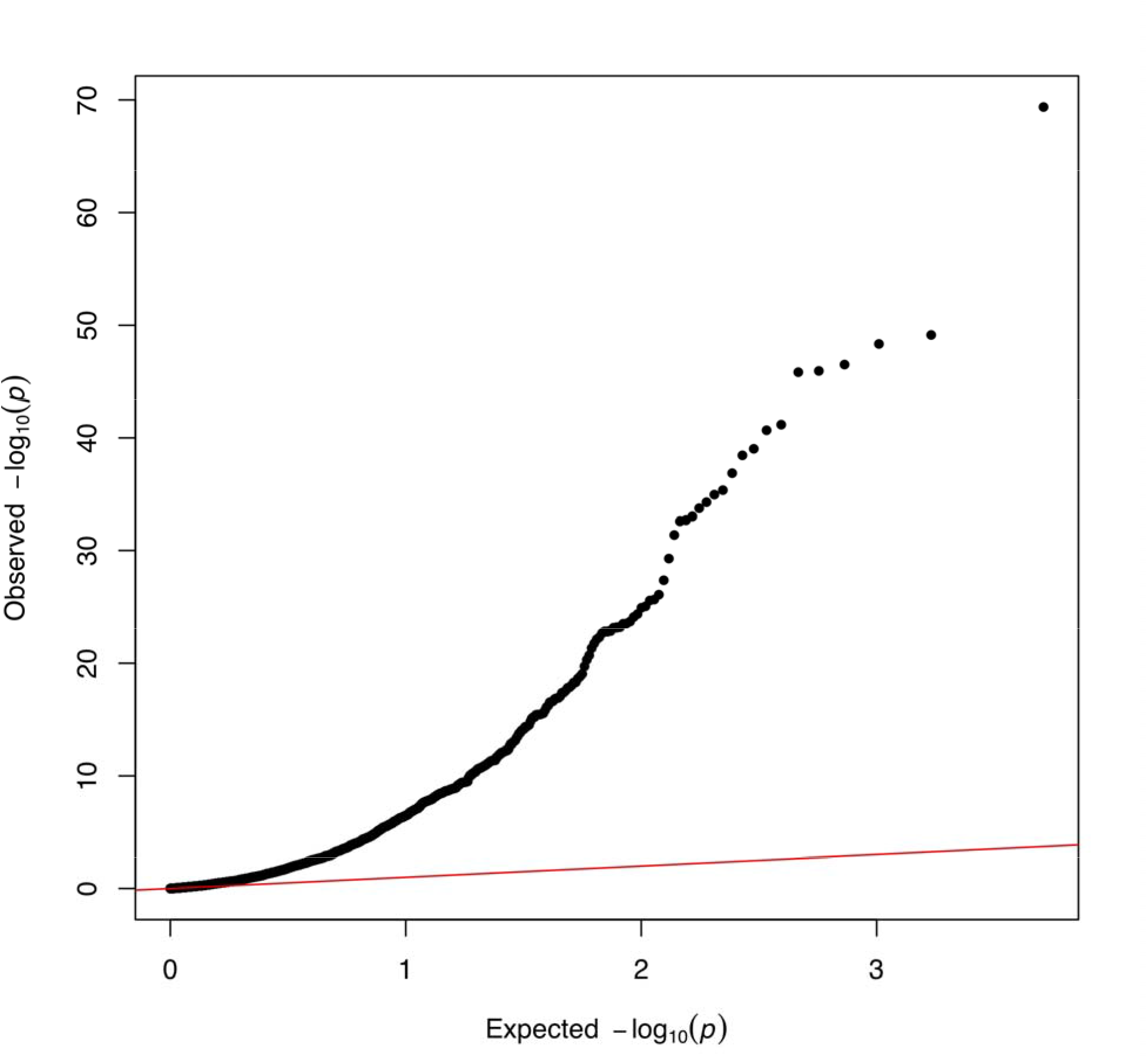
Quantile-Quantile plot of −log10 p-values for factors influencing being tested for COVID-19 in UK Biobank. The x-axis represents the expected p-value for 2,556 hypothesis tests and y-axis represents the observed p-values. The red line represents the expected relationship under the null hypothesis of no associations.

### Sample selection pressures for COVID-19 testing

While some of the factors that impact the sampling processes may be common across all modes of sampling listed above, some will be mode-specific. In general, these factors will differ across national and healthcare system contexts. Here we list a series of possible selection pressures acting upon COVID-19 testing and case identification/definition and detail how they may bias inference if left unexplored.

#### Symptom severity

With few notable exceptions (e.g. (3)), population testing for COVID-19 is not generally performed in random samples. Several countries adopted the strategy of offering tests predominantly to patients experiencing symptoms severe enough to require medical attention, e.g. hospitalisation, as is the case in the UK until the end of April 2020. Many true positive cases in the population will therefore remain undetected and be subject to negative sample selection if enrollment is dependent upon test status. High rates of asymptomatic virus carriers or cases with atypical presentation will further compound this issue.

#### Symptom recognition

Related to but distinct from symptom severity, inclusion in COVID-19 datasets will vary based upon symptom recognition (33). If an individual fails to recognise the correct symptoms or deems their symptoms to be nonsevere, they are less likely to seek medical attention and therefore be tested for COVID-19. People will also assess their symptom severity differently; those with health related anxiety may be more likely to over-report symptoms, while those with less awareness or access to health advice may be under-represented. This problem may be compounded by changing symptom guidelines which could induce systematic relationships between symptom presentation and testing (33,34).

#### Occupation

In many countries, frontline healthcare workers are far more likely to be tested for COVID-19 than the general population (5,35) due to their proximity to the virus and the potential consequences of infection related transmission (36). As such, they will be heavily over-represented in samples conditional on test status. Other key workers may be at high risk of infection due to large numbers of contacts relative to non-key workers, and may therefore be over-represented in samples conditional on test status or cause of death. Any factors related to these occupations (e.g. ethnicity, socio-economic position, age and baseline health) will therefore also be associated with sample selection. **Figure 2B** illustrates an example where the hypothesised risk factor does not need to influence sample selection causally, it could simply be associated due to a confounding between the risk factor and sample selection.

#### Place of residence and social connectedness

A number of more distal or indirect influences on sample selection likely exist. People with better access to healthcare services may be more likely to be tested than those with poorer access. Those in areas with a greater number of medical services or better public transport may find it easier to access services for testing, while those in areas with lower local medical service-utilisation may be more likely to be tested as a function of service capacity (37). People living in areas with stronger spatial or social ties to existing outbreaks may also be more likely to be tested due to increased medical vigilance in those areas. Family and community support networks are also likely to influence access to medical care, for instance, those with caring responsibilities and weak support networks may be less able to seek medical attention (38).

#### Frailty

Some groups of the population, such as elderly in care homes, are treated differently in terms of reporting on COVID-19 in different countries (39). For example in the UK early reports of deaths “due to COVID-19” may have been conflated with deaths “while infected with COVID-19” (40). Individuals at high risk are more likely to be tested in general, but specific demographics at high risk such as those in care homes have been liable to under-representation. A challenge that arises with trying to evaluate the problem of collider bias is that it may be difficult to ascertain if particular groups with COVID-19 are being over or under represented in the selected sample, making sensitivity analysis difficult.

### Sample selection pressures for voluntary self-reporting

Sample selection pressures for voluntary self-reporting COVID-19 efforts are likely distinct from those for COVID-19 testing.

#### Internet access and Technological Engagement

Sample recruitment via internet applications has been shown to under-represent certain groups (26,41). Furthermore, voluntary “pull-in” data collection methods have been shown to produce more engaged but less representative samples than “push out” advertisement methods (27). These groups likely have greater access to electronic methods of data collection, and greater engagement in social media campaigns that are designed to recruit participants. As such, younger people are more likely to be over-represented in app based voluntary participation studies (23).

#### Medical and scientific interest

Voluntary participation studies are likely to contain a disproportionate amount of people who have a strong medical or scientific interest. It is likely that these people will themselves have greater health awareness, healthier behaviour, be more educated, and have higher incomes (25,42).

#### Ethnicity

Some groups may experience barriers into voluntary participation of scientific studies due to many factors, for example language, cultural norms or access to information.

Many of the factors for being tested or being included in datasets described here are borne out in the analysis of the UK Biobank test data (Box 2).

### Sample selection pressures for hospitalisation

Several of the factors described above are also relevant for selection into samples from hospital admissions. Pre-existing medical conditions, obesity, smoking and age are factors that are of interest to understanding the aetiology of COVID-19, but they also strongly associate with hospitalisation. Different ethnic groups or socio-economic strata may have differential hospital admission rates. At the same time, COVID-19 infection and severity likely have an influence on hospitalisation (8-10,28). Hence, studies that are based on data derived from hospitalised patients or records are liable to correlations induced between COVID-19 measures and a wide range of clinical measures that do not reflect causal effects or exist in the general population.

## Methods for overcoming collider bias

In this section we describe methods to either overcome bias or evaluate how sensitive any associations could be to collider bias. The primary task in any analysis is to evaluate the extent to which sample selection is likely to have actually occurred. This can be done by comparing means and prevalences in the selected sample against those obtained from external data that represents the target population. Ideally, this would be done for the hypothesised risk factor and outcome, as well as any related variables. If there are even subtle departures in the characteristics of the study sample from the general population then this provides evidence of selective sampling. With respect to analysis of COVID-19 disease risk, one major obstacle to this endeavour is that in most cases the actual prevalence of infection in the general population is unknown, making it impossible to prove an absence of selection through validation.

If a study is at risk of selective sampling, the unfortunate truth is that it is very difficult to prove that any method has resolved issues with collider bias. Sensitivity analyses are therefore crucial in exploring factors that could be related to selection, and examining robustness of conclusions to plausible selection mechanisms.

Several methods exist that do attempt to adjust for collider bias or examine how sensitive the study is to collider bias. The likelihood and extent of collider bias induced by sample selection can be evaluated by comparing distributions of variables in the sample with those in the target population (or a representative sample of the target population). This provides information about the profile of individuals selected into the sample from the target population of interest, such as whether they tend to be older or more likely to have comorbidities. It is particularly valuable to report these comparisons for key variables in the analysis, such as the hypothesised risk factor and outcome, and other variables related to these.

The applicability of different methods depends on the data that are available on non-participants. These methods can broadly be split into two categories: a) where the selected sample is nested within a larger dataset that comprises samples believed to be representative of the target population, or b) where the entire dataset comprises only the selected samples used for hypothesis testing (stand-alone).

### Nested sample

In the case that we have a selected sample with COVID-19 measures, which is a subset of a sample that is representative of the target population, one approach is to use inverse probability weighting (43,44). Here the causal effect of risk factor on outcome is examined using a weighted regression, where the participants who are overrepresented are down-weighted and the participants who are underrepresented are up-weighted. In practice, we estimate the probability of different individuals selecting into the sample from the population-representative sample based on their measured covariates, using a statistical model (the “sample selection model”), and use this to create a weight for each participant (45). An example is where the study sample is those with a positive covid test, nested within the UK Biobank study. If we assume that UK Biobank is representative of the target population (the general population of the UK), then we can use data from UK Biobank to estimate the probability of having a covid test for each individual in UK Biobank. We can then appropriately re-weight the study sample to represent the population of UK Biobank.

Seaman and White (2013) provide a detailed overview of the practical considerations and assumptions for inverse probability weighting, such as correct specification of the sample selection model, variable selection and approaches for handling unstable weights (i.e. weights which are zero or near-zero). An additional assumption for inverse probability weighting is that each individual in the target population must have a non-zero probability of being selected into the sample. Neither this assumption, nor the assumption that the selection model has been correctly specified, are testable using the observed data. A conceptually related approach, using propensity score matching, is sometimes used to avoid index event bias (46,47).

### Stand-alone samples

When we only have data on the study sample (e.g. only data on participants who were tested for COVID-19) it is not possible to estimate the selection model directly since non-selected (untested) individuals are unobserved. Instead, it is important to apply sensitivity analyses to assess the plausibility that sample selection induces collider bias.

#### Bounds and parameter searches

It is possible to infer the extent of collider bias given knowledge of the likely size and direction of influences of risk factor and outcome on sample selection (whether these are direct, or via other factors) (15,48,49). However, this approach depends on the size and direction being correct, and there being no other factors influencing selection. It is therefore important to explore different possible sample selection mechanisms and examine their impact on study conclusions. We created a simple web application guided by these assumptions to allow researchers to explore simple patterns of selection that would be required to induce an observational association: http://apps.mrcieu.ac.uk/ascrtain/. In **Figure 4** we use a recent report of a protective association of smoking on COVID-19 infection (28) to explore the magnitude of collider bias that can be induced due to selected sampling, under the null hypothesis of no causal effect.

Several other approaches have also been implemented into convenient online web apps (Appendix). For example Smith and VanderWeele (2019) proposed a sensitivity analysis which allows researchers to bound their estimates by specifying sensitivity parameters representing the strength of sample selection (in terms of relative risk ratios). They also provide an ‘E-value’, which is the smallest magnitude of these parameters that would explain away an observed association (50). Aronow and Lee (2013) proposed a sensitivity analysis for sample averages based on inverse probability weighting when the weights cannot be estimated but are assumed to be bounded between two researcher-specified values (51). This work has been generalised to allow regression models to incorporate relevant external information (e.g. summary statistics from the census) (52). Zhao et al (2017) developed a sensitivity analysis for the degree to which estimated probability weights differ from the true probability weights due to misspecification (53). This approach is particularly useful when we can estimate probability weights including some, but not necessarily all, of the relevant predictors of sample selection. For example, where we have the study sample being those with a covid test, nested within the UK Biobank, we may have data within UK Biobank on some predictors of testing, but there could be other factors that are not recorded within UK Biobank (e.g. general predisposition for seeking healthcare).

These sensitivity analysis approaches allow researchers to explore whether there are credible collider structures that could explain away observational associations. However, they do not represent an exhaustive set of models that could give rise to bias, nor do they necessarily prove that collider bias influences the results. If the risk factor for selection is itself the result of further upstream causes then it is important that the impact of these upstream selection effects are considered (i.e. not only how the risk factor influences selection but also how the causes of the risk factor and/or the causes of the outcome influence selection e.g. **Figure 2B**). While these upstream causes may individually have a small effect on selection, it is possible that lots of factors with individually small effects could jointly have a large selection effect and introduce collider bias (Groenwold et al. 2016).

#### Negative control analyses

If there are factors measured in the selected sample that are known to have no influence on the outcome, then testing these factors for association with the outcome within the selected sample can serve as a negative control (54,55). By virtue, negative control associations should be null and they are therefore useful as a tool to provide evidence in support of selection. If we observe associations with larger magnitudes than expected then this indicates that the sample is selected on both the negative control and the outcome of interest (56,57).

#### Correlation analyses

Conceptually similar to the negative controls approach above, when a sample is selected, all the features that influenced selection become correlated within the sample (except for the highly unlikely case that causes are perfectly multiplicative). Testing for correlations amongst hypothesised risk factors where it is expected that there should be no relationship can indicate the presence and magnitude of sampling bias, and therefore the likelihood of collider bias distorting the primary analysis (58).

### Implications

The majority of scientific evidence informing policy and clinical decision making during the COVID-19 pandemic has come from observational studies (59). We have illustrated how these observational studies are particularly susceptible to non-random sampling. Randomised clinical trials will provide experimental evidence for treatment, but experimental studies of infection will not be possible for ethical reasons. The impact of collider bias on inferences from observational studies could be considerable, not only for disease transmission modelling (60,61), but also for causal inference (7) and prediction modelling (2).

While many approaches exist that attempt to ameliorate the problem of collider bias, they rely on untestable assumptions. It is difficult to know the extent of sample selection, and even if that were known it cannot be proven that it has been fully accounted for by any method. Representative population surveys or sampling strategies that avoid the problems of collider bias (62) are urgently required to provide reliable evidence. Results from samples that are likely not representative of the target population should be treated with caution by scientists and policy makers.

## Data Availability

All analysis was performed on UK Biobank data

https://github.com/explodecomputer/covid_ascertainment

## Appendix

Exploring bounds and spaces that could explain an observational association can easily be achieved using a range of packages and apps:

- AscRtain app: http://apps.mrcieu.ac.uk/ascrtain/
- CollideR app (13): https://watzilei.com/shiny/collider/
- Selection bias app (50): https://selection-bias.herokuapp.com/
- Bias app (49): https://remlapmot.shinyapps.io/bias-app/
- Lavaan R package (63): http://lavaan.ugent.be/
- Dagitty R package (64): http://www.dagitty.net/
- simMixedDAG: https://github.com/IyarLin/simMixedDAG

## Acknowledgements

We are grateful to Josephine Walker for helpful comments on this manuscript. This research has been conducted using the UK Biobank Resource under Application Number 16729. The Medical Research Council (MRC) and the University of Bristol support the MRC Integrative Epidemiology Unit [MC_UU_12013/1, MC_UU_12013/9, MC_UU_00011/1]. GG is supported by an ESRC postdoctoral fellowship [ES/T009101/1]. NMD is supported by a Norwegian Research Council Grant number 295989. GH is supported by the Wellcome Trust and Royal Society [208806/Z/17/Z].

## Author contributions

G.H., N.M.D., L.Z. conceived the idea

G.H. performed the analysis

G.G., G.H., and T.P. wrote the software

All authors discussed the results and contributed to the final manuscript

## Patient and public involvement

This research was done without patient involvement. Patients were not invited to comment on the study design and were not consulted to develop patient relevant outcomes or interpret the results. Patients were not invited to contribute to the writing or editing of this document for readability or accuracy.

#### Box 1: Collider bias in the context of prediction and aetiological studies

An aetiological study seeks to identify causes of the outcome of interest (“causal factors”), whereas a predictive study aims to predict the outcome from a range of variables (“predictors”) which need not be causal. The term “risk factor” has been used synonymously for both causal factors and predictors in the literature (65,66).

Risk factors measured in observational studies, may associate with outcomes of interest (e.g. hospitalised with COVID-19), for many reasons. For example, the factor may affect the outcome (true causal interpretation), statistical evidence of association may be purely due to chance, the outcome may affect the factor (reverse causation), there may be a third factor that causes both the exposure and the outcome (confounding), or the exposure and outcome (or causes of the exposure and/or outcome) may influence likelihood of being selected into the study (collider bias).

Aetiological studies are in principle only concerned with the causal effect, and aim to avoid all forms of bias. By contrast, some forms of bias such as confounding or reverse causation can actually improve the performance of a prediction study. As long as the causal structure by which the study sample is drawn from the target population is the same as in the population in which predictions will be made, it can be of benefit to leverage these distinct association mechanisms to improve prediction accuracy (67,68).

Under certain circumstances collider bias can improve prediction performance if the training sample and the sample to be predicted have the same patterns of sample selection. For example, if the factors causing having a test for COVID-19 are the same/similar across the UK, a predictive model for the result being positive that was developed in London will perform well in the North East if those samples are both selected in the same way. However, collider bias is a problem for the generalisability of both causal inference and prediction in the target population when the training sample is selected, because it induces artifactual associations that are idiosyncratic to that dataset. If the intention is to predict COVID-19 status, rather than COVID-19 status conditional on being tested, the prediction will underperform.

While the term ‘risk factor’ can be ambiguous and refer to either a hypothesised causal determinant or a predictor of the disease, we intentionally use it throughout this paper for the sake of brevity as causal inference and prediction analyses both share a vulnerability to the detrimental impacts of collider bias in the COVID-19 context - where typically the selected samples are being used to develop models relevant to the general population.

#### Box 2. The potential association between ACE inhibitors and COVID-19: why sampling bias matters

One research question that has gained attention is whether blood pressure lowering drugs such as ACE inhibitors (ACE-i) and angiotensin-receptor blockers (ARBs), which act on the Renin-Angiotensin-Aldosterone System (RAAS) system, make patients more susceptible to COVID-19 infection (69-73).

Relationships between ACE-i/ARBs and COVID-19 are to be investigated in clinical trials (74,75), but in the meantime have been rapidly investigated through observational studies (76-78). One such recent analysis used data from a UK COVID-19 symptom tracker app (79), which was released in March just before the UK Lockdown policy was implemented to increase social distancing. The app allows members of the public to contribute to research through self-reporting data including demographics, conditions, medications, symptoms and COVID-19 test results. The researchers observed that people reporting ACE-i use were twice as likely to report COVID-19 symptoms, even after adjusting for differences in age, BMI, sex, diabetes, and heart disease (24).

The researchers investigated whether sampling bias may play a role. If taking ACE-i and having COVID-19 symptoms would lead to being either less or more likely to sign up to the app or contribute data, this could induce an association between these factors (**Figure 2A**). Since ACE-is are prescribed to those with diabetes, heart disease, or hypertension, ACE-i users are likely to be considered high-risk for COVID-19 (80). They are therefore potentially more sensitised to their current health status and may be more likely to use the app (81,82). People who are COVID-19 symptomatic may also be more likely to remember to contribute data than asymptomatic people. Taken together, this could result in a false or inflated association between taking ACE-i and COVID-19. However, in reality, deciding in which direction ACE-i and COVID-19 symptoms would influence participation is complicated. For example, people with severe COVID-19 symptoms who are hospitalised could be too ill to contribute data.

Careful consideration is required for each set of exposures and outcomes that are studied. Amongst those participants who were actually tested in the COVID-19 symptom tracker app study, there was no evidence for an association between ACE-i use and COVID-19 positive status (24). In this analysis there are joint selection pressures of a) factors underlying being tested and b) factors underlying app participation.

Should ACE-i use truly increase risk of COVID-19 infection, it could imply that observational results for disease progression studies are influenced by collider bias. For example, it has been reported that ACE-i/ARB use may be protective against severe symptoms, conditional on already being infected (83,84), which is consistent with index event bias as illustrated in **Figure 2C**.

It is important to consider the plausibility of the different selection pathways, both statistically (for example, through methods such as bounds and parameter searches) and biologically. Such considerations will ensure that data interpretation is at least robust to known biases of unknown magnitude, and policy decisions are based on the best interpretation of the scientific evidence. Indeed, in consideration of the benefits that ACE-i/ARBs have on the cardio-respiratory system, current guidelines should continue to recommend use of these drugs until there is sufficiently reliable scientific evidence against this (85,86).

#### Box 3. Factors influencing being tested in UK Biobank

In April 2020, General Practices across the UK released primary care data on COVID-19 testing for linkage to the participants in the UK Biobank project (87,88) and analyses are already appearing (89). Of the 486,967 participants, 1,410 currently have data on COVID-19 testing. While it may be tempting to look for factors that influence whether an individual tests positive, it is crucial to evaluate the potential that those tested are not a random sample of the UK-Biobank participants (who are themselves not a random sample of the UK population).

We examined 2,556 different characteristics for association with whether or not a UK Biobank participant had been tested for COVID-19. There was very large enrichment for associations (Figure 2), with 811 of the phenotypes (32%) giving rise to a false discovery rate < 0.05. These associations involved a wide range of traits, including measures of frailty, medications used, genetic principal components, air pollution, socio-economic status, hypertension and other cardiovascular traits, anthropometric measures, psychological measures, behavioural traits, and nutritional measures. A full list of all traits assessed and their associations with whether a participant had COVID-19 test data are available in Supplementary Table 1. The first genetic principal component, which relates to major ethnic groups, was one of the strongest associations with being tested, which may have implications for understanding the association of race on testing positive for COVID-19 (89).

We can not know the actual COVID-19 prevalence amongst all participants, but if it is different from the prevalence amongst those tested, then every one of the traits listed above could be associated with COVID-19 in the dataset solely due to collider bias, or at least the magnitude of those associations could be biased as a result. The fact that the UK Biobank data are already a non-random sample of the UK population further complicates the matter (30).

Ideally, inverse-probability weighted regressions would be performed to minimise any such bias. However, because we can not know the COVID-19 status of participants outside the tested group (sampling fractions), such weights will be impossible to calculate without strong assumptions that are currently untestable (53). Inverse-probability-weighting also depends on the selection model being correctly specified, including that all characteristics predicting selection (that are related to variables in the analysis model) have been included, and in the right functional form. As with unmeasured confounding, there is always the possibility of having unmeasured selection factors.

##### Methods

UK-Biobank phenotypes were processed using the PHESANT pipeline (90) and filtered to include only quantitative traits or case-control traits that had at least 10,000 cases. In addition, sex, genotype chip and the first 40 genetic principal components were included for analysis (2,556 traits in total). A ‘tested’ variable was generated that indicated whether an individual had been tested for COVID-19 or not within UK Biobank, and logistic regression was performed for each of the 2,556 traits against the ‘tested’ variable. Code: https://github.com/explodecomputer/covid_ascertainment

**Figure 3:**
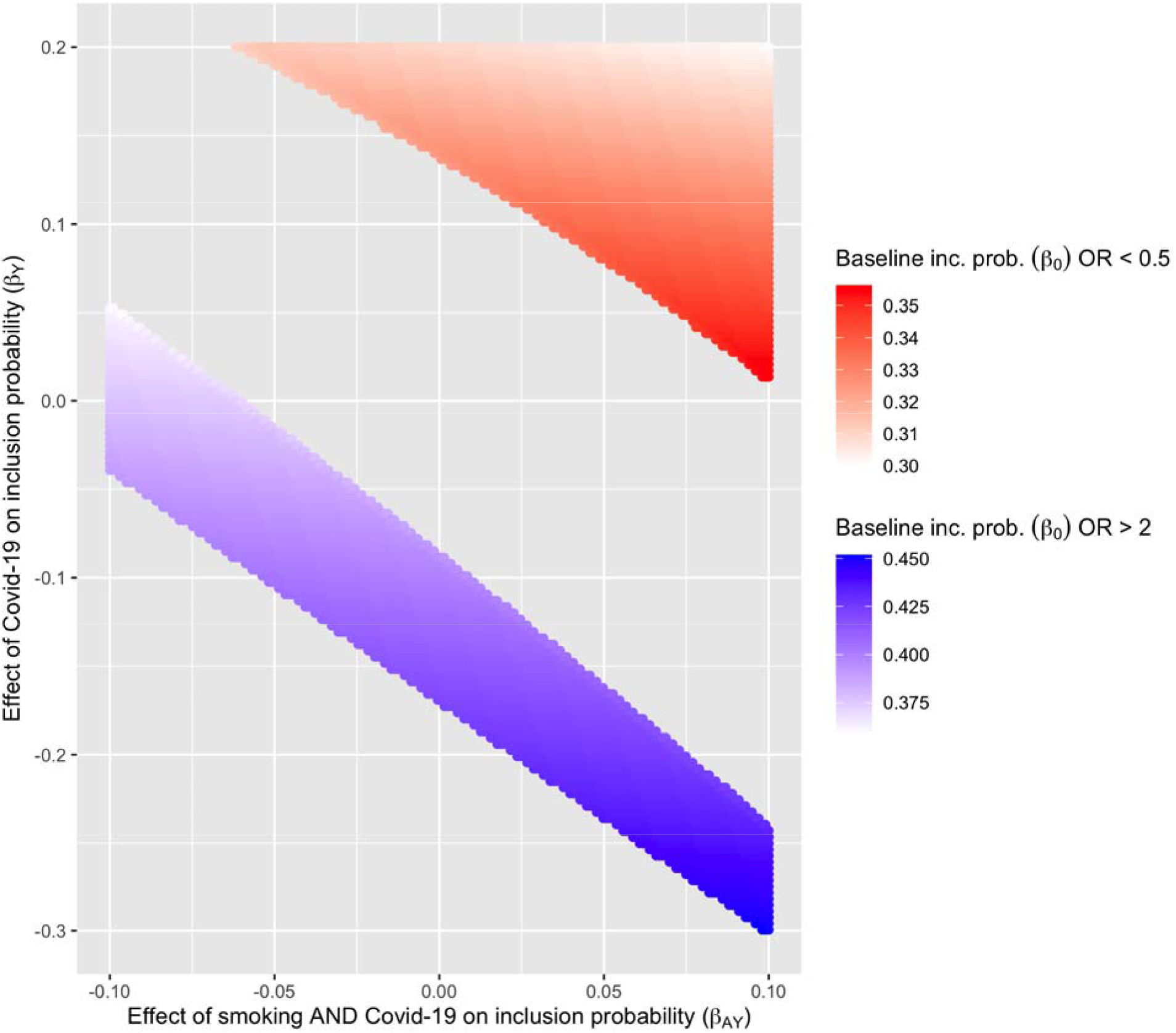
Large cassociations can be induced by collider bias under the null hypothesis of no causal relationship, using scenarios similar to those reported for the observed protective association of smoking on COVID-19 infection. Assume a simple scenario in which the hypothesised exposure (A) and outcome (Y) are both binary and each influence probability of being selected into the sample (S) e.g. where is the baseline probability of being selected, is the effect of A, is the effect of Y and is the effect of the interaction between A and Y. The selection mechanism in question is represented in Figure 1B (without the interaction term drawn). This plot shows which combinations of these parameters would be required to induce an apparent risk effect with magnitude OR > 2 (blue region) or an apparent protective effect with magnitude OR < 0.5 (red region) under the null hypothesis of no causal effect (49). To create a simplified scenario similar to that in Miyara et al 2020 we use a general population prevalence of smoking of 0.27 and a sample prevalence of 0.05, thus fixing *β_A_* at 0.22. Because the prevalence of COVID-19 is not known in the general population, we allow the sample to be over or under representative (y-axis). We also allow modest interaction effects. Calculating over this parameter space, 40% of all possible combinations lead to an artifactual 2-fold protective or risk association operating through this simple model of bias alone. It is important to disclose this level of uncertainty when publishing observational estimates.

## Supplementary tables

**Supplementary Table 1:** Association results for each of 2,556 variables in the UK Biobank cohort, testing for their influence on being tested for COVID-19

